# Oxygen-based endotypes of Obstructive Sleep Apnea

**DOI:** 10.64898/2026.06.03.26354835

**Authors:** Andrew Wellman, Ludovico Messineo, Ali Azarbarzin, Neda Esmaeili, Atqiya Aishah, Daniel Vena, Jeffrey Sumner, David P White, Scott A Sands

## Abstract

**Objective:** Several endotypes contribute to the development of Obstructive Sleep Apnea (OSA). However, efforts to measure these endotypes have been challenging. In this paper, we propose a new method that overcomes some of these challenges.

**Methods:** To test the feasibility of this new method, data from the Sleep Heart Health Study (SHHS) were analyzed and two oxygen-based endotypes were identified and plotted on a graphical model: the steady-state SpO2 and the SpO2 arousal threshold. The first is the oxygen saturation that would occur during sleep if there were no arousals, and it is a measure of upper airway collapsibility (a more collapsible airway produces a lower SpO2). The latter is the oxygen saturation that triggers arousals. These endotypes were validated by assessing their ability to detect positional and state-related changes in airway collapsibility and arousal threshold.

**Results:** The study showed that it was feasible to measure oxygen-based endotypes in 95% of SHHS participants. As expected, steady-state SpO2 was lower during supine vs. non-supine sleep, as well as during REM vs. NREM sleep. Also, the SpO2 arousal threshold was similar between supine and non-supine sleep. However, SpO2 arousal threshold was not lower in REM sleep vs. NREM sleep. Therefore, in 3 of the 4 conditions, the oxygen-based endotypes moved in the expected direction due to positional or sleep state changes.

**Conclusion:** Although further validation experiments are required, this study indicates that OSA endotyping using the pulse oximetry signal is feasible. The oxygen-based endotypes could be used to aid therapeutic decision making.

## INTRODUCTION

The development of Obstructive Sleep Apnea (OSA) is influenced by various endotypes, such as a collapsible upper airway, an oversensitive ventilatory control system, poor pharyngeal muscle responsiveness during sleep, and a low respiratory arousal threshold (1, 2). Knowing these endotypes could be useful for classifying patients or choosing therapy. However, measuring the endotypes is difficult. In our early studies, we developed the CPAP drop method to measure the endotypes (3, 4). This approach involved reducing CPAP to elicit ventilatory disturbances. The responses to these disturbances were analyzed using a dynamic breathing model to infer endotypes. The method provided not only estimates of the endotypes, but also a graphical model illustrating the interaction of endotypes in OSA pathogenesis. However, it faced notable limitations: the need for specialized equipment and personnel, complexity of the mathematical modeling, and its impracticality in clinical settings.

To address these shortcomings, we designed algorithms to extract endotypes from standard polysomnography (PSG) (5–7). This adaptation allowed endotype determination without specialized equipment and expanded its utility to existing PSG datasets. Nevertheless, this method still presents some challenges. It relies on the presence of apneas and hypopneas to disturb ventilation, thus not accounting for endotype assessment during stable breathing. Also, it lacks the visual model available with the CPAP drop method, an aspect that could be important for choosing specific therapies. Furthermore, it is constrained by the need for high-quality PSG data and thus is not readily applicable to home-based studies.

The novel approach presented in this paper addresses several of these issues. In particular, it determines the endotypes from the pulse oximetry signal, which is easy to acquire and could be performed over multiple nights in a home setting. Furthermore, it simplifies interpretation by circumventing the need for complex mathematical modeling. Notably, this technique allows for endotype assessment during both disturbed and stable breathing episodes, providing a more comprehensive estimate of the endotypes. Finally, it reintroduces the graphical model for understanding endotype interactions, enhancing its potential for clinical application.

To validate the oxygen-based endotyping method, several hypotheses were tested in the Sleep Heart Health Study (SHHS) dataset (8, 9). First, we examined whether it would be feasible to estimate the oxygen-based endotypes in this cohort. Second, we determined if the endotypes moved in the expected direction following positional and sleep stage changes. And third, we explored whether the graphical model of the endotypes agreed with various measures of OSA severity.

## METHODS

### SpO2 Arousal Threshold

To explain the oxygen-based endotyping method, the idealized SpO2 tracing in Figure 1 will be used (where SpO2 serves as an estimate of SaO2). This method requires the measurement of awake SpO2 in the recumbent position, which is assumed to be 96% at time 0 in Figure 1. As sleep begins (time > 0 minutes), the upper airway narrows, limiting ventilation and causing a drop in oxygen saturation. For the first 17 minutes of this recording, the patient’s airway remains sufficiently patent to maintain a stable SpO2 of 91%. However, at the 17-minute mark, the SpO2 begins to decrease again, likely due to increased airway obstruction, possibly from a change in position or sleep stage. Regardless of the cause, the SpO2 continues to decline until the airway abruptly opens at 19 minutes, leading to a sharp increase in SpO2. It is assumed that this airway opening is triggered by an arousal. While the airway can reopen without an arousal (10), in both scenarios the end result is the cessation of an apnea or hypopnea, which is what we are interested in capturing.

**Figure 1.**
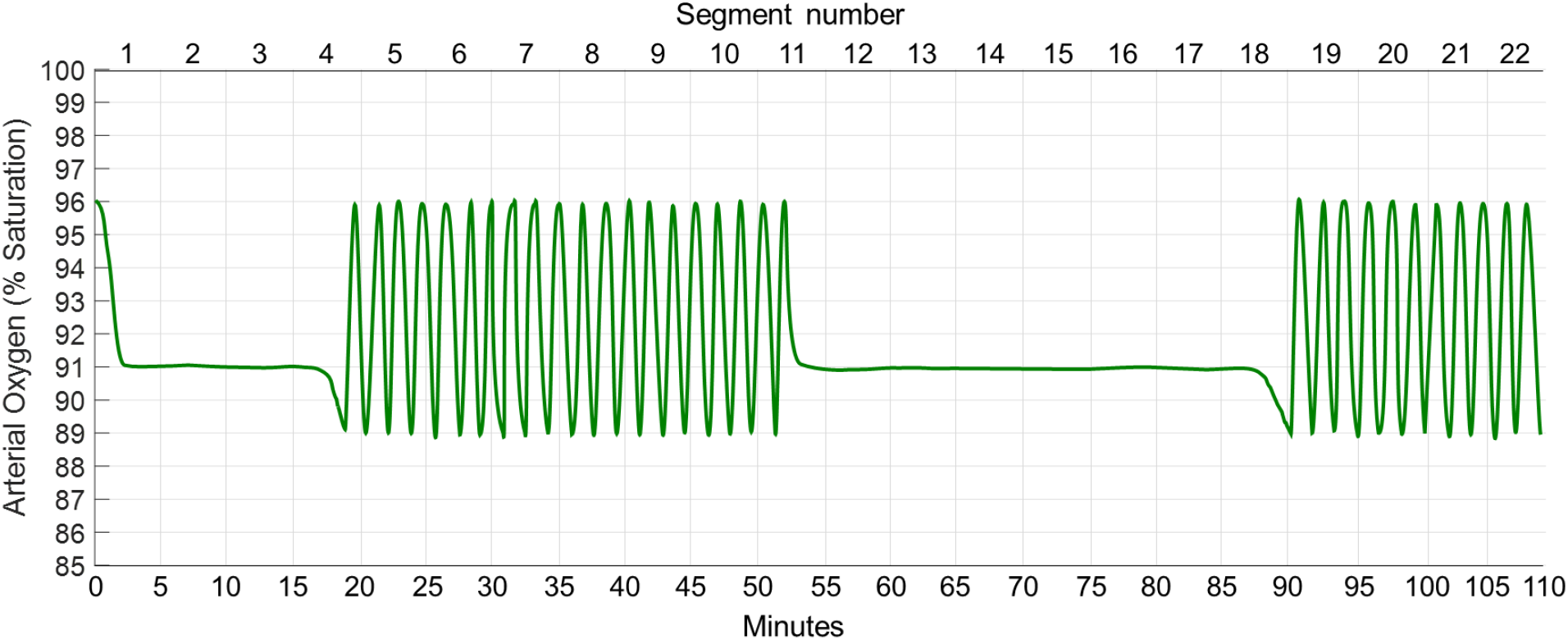
Drawing of an SpO2 signal during sleep in a person with OSA.

Following the sharp increase and return to baseline (time = 20 minutes), the SpO2 falls again as the patient goes back to sleep. When the SpO2 reaches 89%, another arousal occurs, leading to an increase in SpO2, and so on. The patient is having cyclic OSA from time 17-52 minutes. In this method, it is assumed that cycling is driven by upper airway obstruction and not ventilatory control instability. If ventilatory control instability is thought to be the main cause of cycling, then this method would not be applicable.

From this figure, it is clear that the patient arouses whenever SpO2 drops to 89%, indicating an SpO2 arousal threshold of 89%. While arousal threshold can be observed during respiratory events, it cannot be directly observed during periods of stable breathing when there are no respiratory events. However, it can still be partially observed during these stable breathing periods. For instance, it can be inferred that the SpO2 arousal threshold is below 91% between minutes 2 through 17 and 54 through 87 in Figure 1. If it were above 91%, then arousals would occur and there would be no stable breathing. Thus, during periods of stable breathing, the SpO2 arousal threshold is below the observed stable-breathing SpO2. On the other hand, during periods of unstable breathing, the arousal threshold is equal to the nadir SpO2 values. These two pieces of information can be used to estimate the arousal threshold for the entire night, as described below.

#### Definitions of stable and unstable breathing

Before describing how the SpO2 arousal threshold is estimated, it is necessary to define stable and unstable breathing. To do this, the study is divided into 5-minute non-overlapping segments. Segments are labeled unstable if they contain at least one respiratory event. A respiratory event is defined the same as it is for the oxygen desaturation index (≥ 3% drop in SpO2). This definition of an “unstable segment” is arbitrary, but it was chosen because it corresponds to an apnea-hypopnea index (AHI) of 12/hr (if there were one event every 5 minutes), which is mild OSA. A segment is stable if there are no respiratory events within it. This, too, is arbitrary, but it was chosen because 5 minutes is probably the minimum time needed to develop a quasi-steady state. In Figure 1, segments 1-3 and 12-18 are stable, whereas segments 5-11 and 19-22 are unstable.

#### Estimation of the SpO2 arousal threshold

To determine the SpO2 arousal threshold, we first create a table listing the asleep segments (Table 1). Each segment is categorized as either stable or unstable based on the definitions provided above. For unstable segments, the average of the SpO2 nadirs is calculated and denoted with an “equal to” sign, indicating that the SpO2 arousal threshold was directly observed in these segments. In contrast, for stable segments, the average SpO2 across the entire segment is calculated and marked with a “less than” sign, signifying that the arousal threshold is lower than the stable breathing SpO2 in these segments. When a dataset includes both “equal to” and “less than” measurements, survival analysis, e.g., Kaplan-Meir method, can be used to make estimates. For the data presented in Table 1, the median SpO2 arousal threshold is estimated to be 89%.

**Table 1.** Data from Figure 1 used to estimate the SpO2 arousal threshold.

| Segment | Label | SpO<sub>2</sub> | = or < | Segment | Label | SpO<sub>2</sub> | = or < |
| --- | --- | --- | --- | --- | --- | --- | --- |
| 1 | stable | 91.8 | < | 12 | stable | 91 | < |
| 2 | stable | 91 | < | 13 | stable | 91 | < |
| 3 | stable | 91 | < | 14 | stable | 91 | < |
| 4 | unstable | 89 | = | 15 | stable | 91 | < |
| 5 | unstable | 89 | = | 16 | stable | 91 | < |
| 6 | unstable | 89 | = | 17 | stable | 91 | < |
| 7 | unstable | 89 | = | 18 | stable | 90.5 | < |
| 8 | unstable | 89 | = | 19 | unstable | 89 | = |
| 9 | unstable | 89 | = | 20 | unstable | 89 | = |
| 10 | unstable | 89 | = | 21 | unstable | 89 | = |
| 11 | unstable | 89 | = | 22 | unstable | 89 | = |

#### Interpretation of the SpO2 arousal threshold

The *SpO2* arousal threshold in this method differs from the *ventilatory drive* arousal threshold described in previous endotyping papers (3, 4, 6). This difference is illustrated in Figure 2, which shows that as SpO2 decreases, ventilatory drive increases. If the controller gain, or chemosensitivity, is high (thin line), then more ventilatory drive is generated for a given drop in SpO2. As a result, arousal occurs at a higher SpO2 level. On the other hand, if the controller gain is low (bold line), then arousal occurs at a lower SpO2 level. Therefore, the SpO2 arousal threshold differs from the ventilatory drive arousal threshold depending on the controller gain. This difference should be considered when comparing the two metrics.

**Figure 2.**
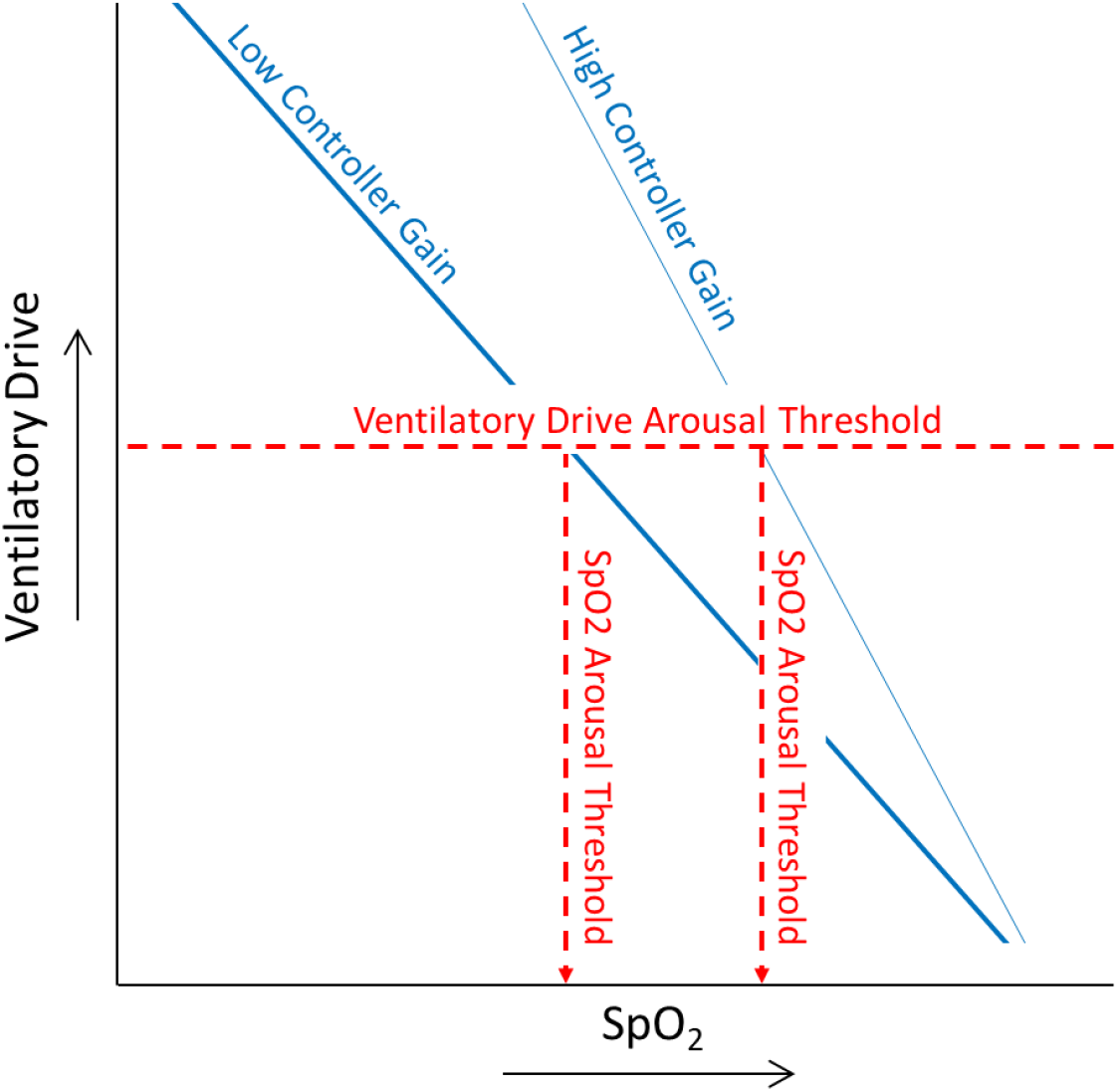
For a given ventilatory drive arousal threshold, a higher controller gain is associated with a higher SpO2 arousal threshold.

### Steady-state SpO2 during sleep

In this section, we argue that the primary cause of the decrease in oxygen saturation during sleep is upper airway obstruction. Consequently, we believe SpO2 can be used to gauge upper airway collapsibility, but only when it is in a relatively steady state, as explained below.

Among the five causes of hypoxemia, hypoventilation is the most likely explanation for the decrease in SpO2 observed during sleep. Low barometric pressure and diffusion limitation do not change from wakefulness to sleep, eliminating them as potential causes. Ventilation-perfusion (V/Q) mismatch and shunt, however, could change due to postural or lung volume-related compression of small airways. However, if the awake SpO2 is measured in the recumbent position, as suggested above, then posture is not changing and thus would not affect V/Q mismatch or shunt. Regarding lung volume, studies show that functional residual capacity decreases by approximately 300 ml from supine wakefulness to sleep (11–14), which would be expected to lower PaO2 by 5-10 mmHg and SaO2 by less than 1% (15). Therefore, V/Q mismatch and shunt are unlikely to account for the observed drop in oxygen saturation during sleep, leaving hypoventilation as the most likely explanation.

Hypoventilation during sleep results from both a reduction in ventilatory drive and upper airway obstruction. However, studies indicate that the sleep-related reduction in ventilatory drive lowers SaO2 by 1% or less (16–19). On the other hand, upper airway obstruction, particularly in patients with OSA, can lower oxygen saturation considerably. For this reason, we believe oxygen saturation can be used as a marker of upper airway obstruction. To do this, however, it is necessary that SpO2 be allowed to equilibrate with ventilation, i.e., reach a relatively steady state. This is generally possible during stable breathing periods when there are no respiratory events. It is not possible, however, when breathing is unstable. Nevertheless, with survival analysis it is possible to make approximations, similar to what was done with arousal threshold estimations.

Figure 3 demonstrates how to determine the steady-state SpO2. This figure contains the same drawing as Figure 1 but with an additional signal added on top of it. The solid line is the actual SpO2, and the dotted line is the steady-state SpO2 that the patient could theoretically achieve if there were no respiratory events. The task is to estimate the dotted line using the solid line.

**Figure 3.**
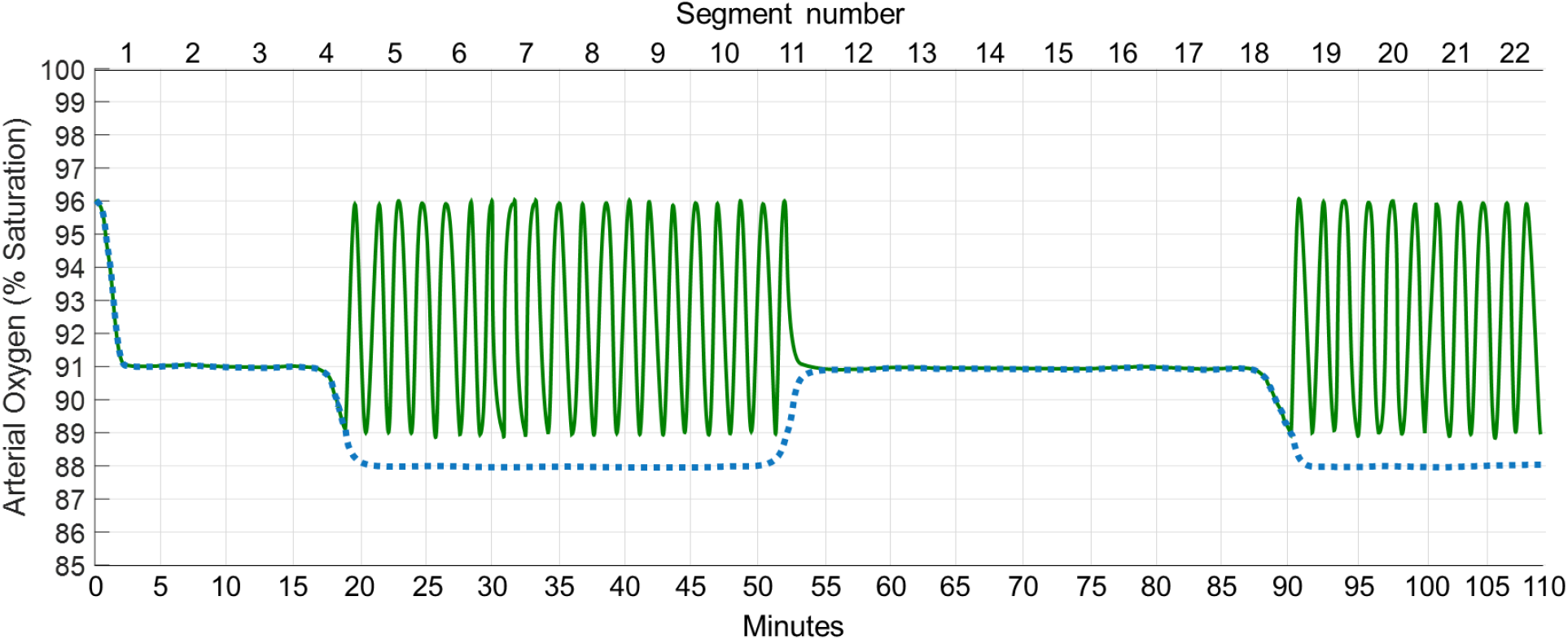
Drawing of an SpO2 signal in a person with OSA. The solid green line is the observed SpO2, and the dotted blue line is the theoretical steady-state SpO2 that the patient could achieve during sleep. Note that the two lines overlay one another during stable breathing, but that the dotted line is below the solid line during unstable breathing.

Like Figure 1, minutes 2 through 17 in Figure 3 show a stable SpO2 of 91%. During periods of stable breathing like this, the observed SpO2 is equal to the steady-state SpO2, i.e., the steady-state SpO2 can be directly inferred from the observed SpO2. However, during periods of unstable breathing (minutes 17 through 52), the steady-state SpO2 cannot be directly observed. Nevertheless, we know that it must be at least below 89% during minutes 17-52; if it were above 89%, then arousals would not be triggered and stable breathing would have occurred. Later in the tracing (time = 54 minutes), the steady-state SpO2 once again increases above the arousal threshold. The most likely reason is a change in airway patency due to a change in position or sleep stage. Regardless of the cause, the steady-state SpO2 is once again observable during this stable breathing episode (minutes 54 to 87). And finally, towards the end of the tracing, the patient is no longer capable of achieving a sustained SpO2 above the arousal threshold, and obstructive events occur. Thus, we can say that the steady-state SpO2 during this period of time (minutes 90 to 110) is < 89%. This information can be used to estimate the steady-state SpO2.

#### Estimation of the steady-state SpO2

Like the SpO2 arousal threshold, the steady-state SpO2 can be estimated using survival analysis approaches. As listed in the previous paragraph, segments 1-3 and 12-18 have observable values equal to approximately 91%, whereas segments 4-11 and 19-22 have indirectly observable values that are < 89%. From these data, the steady-state SpO2 is estimated to be 89%.

#### Interpretation of the steady-state SpO2

The steady-state SpO2 can be interpreted as a measure of airway patency; a more patent airway produces a higher steady-state SpO2, whereas a more obstructed airway produces a lower steady-state SpO2. However, it also contains information about the plant gain for SpO2, which is the amount that SpO2 changes for a given change in ventilation. A high plant gain will produce a larger drop in SpO2 for a 28given reduction in ventilation. Therefore, it is important to understand how plant gain affects the steady-state SpO2 measurement. This is described in further detail in the Discussion.

### SpO2 endotype model of OSA

As mentioned in the Introduction, the oxygen-based endotyping method yields a model that illustrates how the endotypes interact to produce OSA. In the model, the awake SpO2, steady-state SpO2, and SpO2 arousal threshold are plotted on the same axis. Figure 4A shows the model for the data in Figure 1.

**Figure 4.**
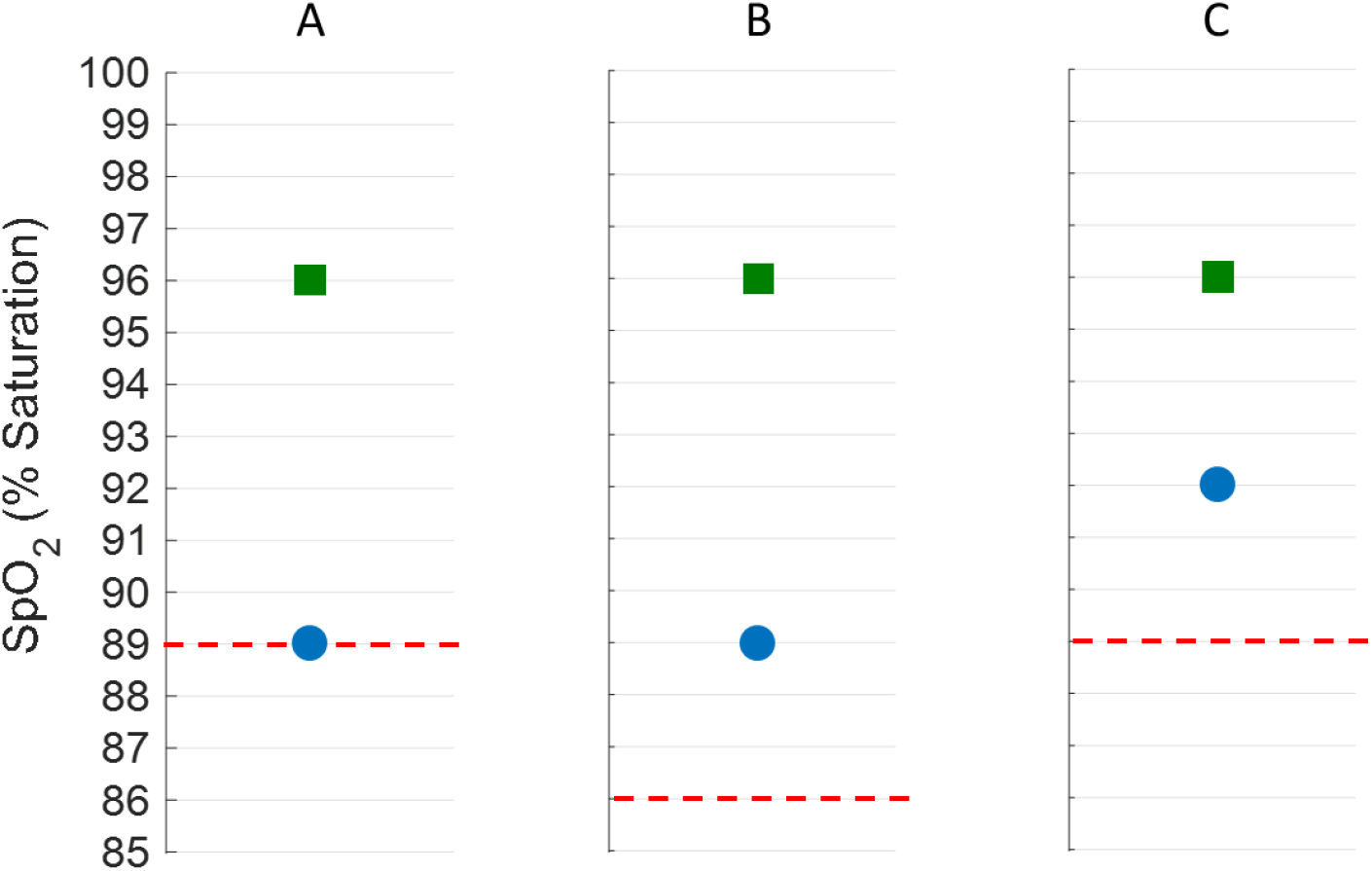
SpO2 endotype model that illustrates how the awake SpO2 (green square), steady-state SpO2 (blue dot), and the arousal threshold (dashed red line) relate to one another. A) Endotype model for the data in Figure 1. B) Hypothetical effect of lowering the SpO2 arousal threshold, as might occur with a hypnotic. C) Effect of raising the steady-state SpO2, as might occur with agents that improve airway patency.

Several insights can be derived from the model. First, the awake SpO2 (green square) is 96%, suggesting that there is no preexisting hypoxemia that might confound interpretation of the oxygen-derived endotypes. Second, the steady-state SpO2 (blue dot) is 7% below wakefulness, indicating that the airway is not severely collapsible. Third, the SpO2 arousal threshold (dashed red line) of 89% is probably appropriate, since it prevents significant hypoxemia from occurring. Fourth, the steady-state SpO2 is close to the SpO2 arousal threshold. In fact, they are equal to one another in this idealized example. This means that small variations in either the steady-state SpO2 or the SpO2 arousal threshold could move breathing into the stable region, as described in the next paragraph.

From this model, treatment could be chosen accordingly. One potential treatment is hypnotic therapy to lower the SpO2 arousal threshold, as depicted in Figure 4B. While the effect size of hypnotics on SpO2 arousal threshold is not known, it is assumed to be a 3% reduction for illustrative purposes. This reduction brings the arousal threshold below the steady-state SpO2, which should eliminate OSA because SpO2 can now be maintained above the level that triggers arousal. However, this treatment runs the risk of allowing sustained hypoxemia (at 89%) to occur. Therefore, lowering the arousal threshold might not be the optimal treatment. An alternative strategy is to raise the steady-state SpO2 (Figure 4C), which could be achieved by reducing airway collapsibility through interventions such as oral appliances or surgery. Depending on the effect size of the selected treatment, this may move the blue dot far enough above the red line to eliminate OSA. In this way, the model can be used to inform therapy. Future studies are needed to determine the effect size of specific treatments on the endotypes.

### Specific methods related to the current study

In the current study, the pulse oximetry-based endotyping method was applied to individuals from the SHHS (8, 9) whose data were accessible through the National Sleep Research Resource (NSRR) (https://sleepdata.org/). Oxygen desaturations were identified using an automated algorithm that detects local minima exhibiting a ≥ 3% drop in SpO2 followed by a ≥ 3% recovery (20). The study was broken into 5-minute, nonoverlapping segments, as described above (see Figure 1). The segments were labeled wake if more than 50% of the 30-second epochs within the segment were scored as wake. Otherwise, they were labeled sleep. Sleep segments were further subdivided into NREM and REM sleep based on the majority of time spent in each stage. The same was done for supine and nonsupine sleep. Asleep segments were labeled stable if there were no desaturations detected in the segment. Otherwise, it was labeled unstable.

The awake SpO2 was computed as the median of the SpO2 in the awake segments. For future applications where EEG is not available, we anticipate that actigraphy could also be used to identify periods of wakefulness. The steady-state SpO2 and SpO2 arousal threshold were calculated for each stage and position using survival analysis approaches, such as the Kaplan-Meier method. The endotypes in the different positions and sleep stages were compared using a Wilcoxon signed-rank test. Spearman correlation analysis was used to compare parameters derived from the model with various measures of OSA severity.

## RESULTS

5,792 SHHS records from the NSRR were analyzed. The participants were 63 ± 11 years old and had a BMI of 28 ± 5 kg/m^2^. Approximately half (52%) were female, and the racial composition was 9% Black, 7% Other, and 85% White. 207 people had underlying hypoxemia, defined as a resting awake SpO2 < 92%. Since pre-existing hypoxemia could confound interpretation of the oxygen-derived endotypes (see Plant Gain for SaO2 in the Discussion), these individuals were excluded from the analysis. An additional 70 people had central sleep apnea, defined as more than 25% of events being central, and were therefore also excluded from the analysis. In total, the endotypes could be determined in 95% of participants in the SHHS.

### Representative results

Typical examples of the endotype model for individuals in the SHHS are presented in Figure 5. An interpretation of these models, along with conjectures regarding potential treatments, is provided. Since these treatments are still unproven, the discussion is speculative, but it is provided here as an illustration of how the model could guide future therapeutic strategies.

**Figure 5.**
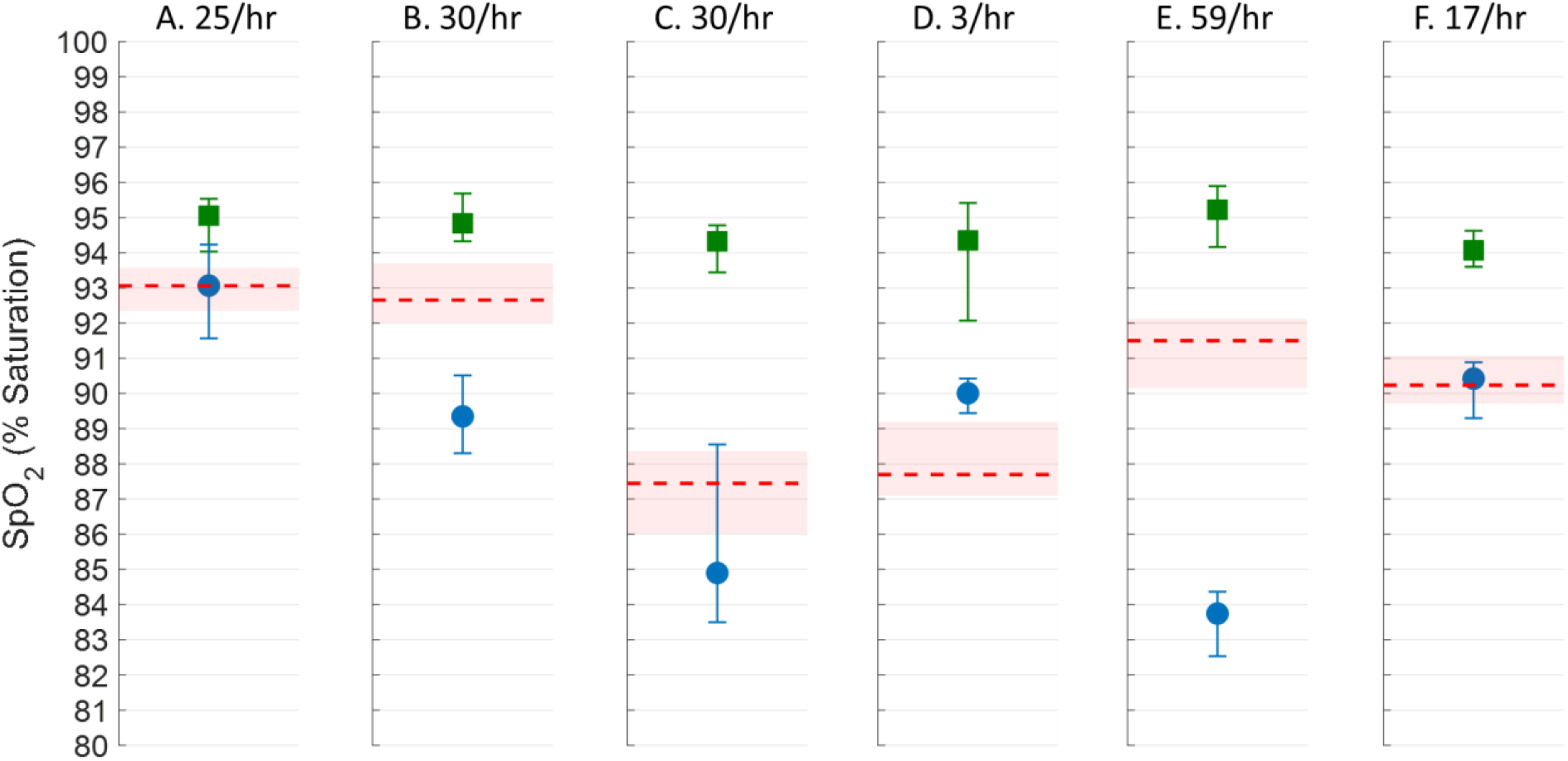
Representative examples of the endotype model for patients in the SHHS. See text for description. Green square – Awake SpO2; Blue dot – Steady-state SpO2; Dashed red line – SpO2 Arousal Threshold.

Figure 5A shows an individual with a minimally collapsible upper airway, as indicated by a steady-state SpO2 that is only two percentage points below the awake SpO2. Despite this, there is moderate OSA based on an ODI of 25/hr. The main problem in this person is that arousals occur, on average, when the oxygen saturation drops to 93%. Therefore, a potential treatment would be through a hypnotic agent to lower the SpO2 arousal threshold.

The subject in Figure 5B also arouses easily at an SpO2 of 92.7%. However, hypnotic therapy alone is probably not appropriate because the steady-state SpO2 is below 90%, and thus lowering the SpO2 arousal threshold might allow for prolonged hypoxemia. If a hypnotic is used in this person, it might need to be combined with a treatment that raises the steady-state SpO2, such oral appliance therapy or surgery.

Unlike Subjects A and B, Subject C does not arouse too easily to respiratory stimuli, since awakening does not occur until the SpO2 decreases below 88%. In this case, hypnotic therapy may not be the most suitable option. Instead, increasing the steady-state SpO2 by improving airway patency would be beneficial. Such treatments could potentially convert Subject C into someone like Subject D. Subject D has an ODI of only 3/hr and is capable of maintaining a stable oxygen saturation well above the level that triggers arousal.

In Subject E, there is a large gap between the steady-state SpO2 and the arousal threshold. For this patient to achieve stable breathing, the blue dot would need to be increased by about 9 percentage points, a sizeable gap that may be difficult to overcome without CPAP. At the other end of the spectrum is Subject F who has a minimal gap. In this person, small adjustments in either the arousal threshold or the steady-state SpO2 could possibly eliminate OSA. Such small changes might be easier to achieve with non-CPAP therapies. Note, however, that the endotypes in Subject F hover around 90%, making hypnotic therapy potentially problematic. Therefore, it may be important to also know the absolute values—and not simply the gap—when choosing therapy. These representative examples illustrate the potential usefulness of the model.

### Supine versus nonsupine sleep

In addition to testing the feasibility of measuring the oxygen-based endotypes, this study also tested the hypothesis that the steady-state SpO2 would be lower in supine sleep compared to nonsupine sleep. This hypothesis was based on the notion that upper airway patency, the main determinant of the steady-state SpO2 endotype, is worse in the supine position. To test this hypothesis, individuals in the SHHS who had an ODI ≥ 10/hr and more than one hour each of supine and nonsupine sleep were identified (n = 1425). In these people, the steady-state SpO2 was 3.1% (IQR 6.8) lower in the supine position than in the nonsupine position (p < 0.001), suggesting that this endotype is capturing the known positional change in airway collapsibility.

Regarding the SpO2 arousal threshold, it was hypothesized that there would be no difference between supine and nonsupine sleep, reflecting the belief that positional changes do not affect arousal mechanisms. Indeed, there was only a 0.3% (IQR 1.5) positional difference in SpO2 arousal threshold (p < 0.001). While the difference is statistically significant, it is small and not clinically important.

### NREM versus REM sleep

Along with testing the effect of position on the endotypes, comparisons were also made between NREM and REM sleep. It was hypothesized that the steady-state SpO2 would be lower in REM sleep, since this stage is associated with more profound airway obstruction. There were 2092 participants in the SHHS who had an ODI ≥ 10/hr and more than one hour each of NREM and REM sleep. The steady-state SpO2 in this group was 4.1% (IQR 6.6) lower in REM sleep compared to NREM sleep (p < 0.001), which is consistent with the known worsening of airway collapsibility in this sleep stage.

For the SpO2 arousal threshold, we postulated that it would be lower in REM sleep because the ventilatory response to chemical stimuli (controller gain) is lower in REM (21–23). However, contrary to our hypothesis, the SpO2 arousal threshold was not different between NREM and REM sleep. The difference was only 0.3% (IQR 1.8) (p < 0.001). Therefore, people aroused at the same level of SpO2 in both NREM and REM sleep. This result is explored in greater detail within the Discussion section.

### Validation of the graphical model

As a test of the model’s accuracy, we examined the correlation between OSA severity and “the gap”—the difference between SpO2 arousal threshold and steady-state SpO2. Theoretically, a large gap should be associated with more severe OSA. This is illustrated in Figure 5E, which shows a large gap in a patient with severe OSA. On the other hand, the person in Figure 5D has a negative gap—the blue dot is above the dashed line—and consequently there is no OSA. To evaluate this theory, the correlation coefficient, *ρ*, between the gap and several indicators of OSA severity was calculated. We found a positive correlation between the gap and the ODI (*ρ* = 0.83, p < 0.001), indicating that a larger gap was associated with a higher ODI. Similarly, the gap’s correlation with AHI, defined using the 3a criteria, was also positive (*ρ* = 0.74, p < 0.001). And finally, the correlation between the gap and the percent stable breathing was *ρ* = −0.88 (p < 0.001), indicating that a larger gap was associated with fewer episodes of stable breathing. This correlation between the gap and OSA severity, which was consistent across various severity measures, lends empirical support to the physiological information provided by the model. It is worth mentioning that our intention is not to replace the AHI with the gap. Rather, the correlation studies were conducted here simply as a validation step.

## DISCUSSION

In this study, two pulse oximetry-derived endotypes were measured, the steady-state SpO2 and the SpO2 arousal threshold. The steady-state SpO2 is the oxygen saturation that would occur during sleep if there were no arousals, and it is a measure of upper airway collapsibility. The SpO2 arousal threshold measures how easily a person arouses to respiratory stimuli. These endotypes, together with the awake SpO2, can be plotted on the same axis to illustrate their interaction in the pathogenesis of OSA. When the steady-state SpO2 is above the arousal threshold, stable breathing is predicted. Conversely, when the steady-state SpO2 is below the arousal threshold, OSA is expected. Furthermore, the greater the difference between the steady-state SpO2 and the arousal threshold, the larger the gap that must be overcome to eliminate OSA. This model could be used to classify individuals or make treatment decisions.

The major finding in this study is that it is feasible to measure the oxygen-based endotypes in a cohort of community-based individuals. Estimates could be obtained in all but 5% of participants. This finding underscores the practicality of our method in assessing endotypes across a broad range of OSA severities.

The endotypes were validated by assessing directional changes in response to alterations in body position and sleep stage. It was found that the endotypes moved in the correct direction in three out of the four conditions. In the fourth condition, REM sleep, we did not find a lower SpO2 arousal threshold, as originally anticipated. While this result reduces confidence in our arousal threshold metric, it also challenges our understanding of how arousals are influenced by REM sleep. We anticipated that the lower controller gain in REM sleep (21–23) would reduce the SpO2 at which arousal occurred in this sleep stage. However, recent data suggest that people arouse at a lower level of ventilatory drive during REM sleep (24), which could offset the low controller gain and cause arousal to occur at similar oxygen saturations across NREM and REM sleep. Before drawing these conclusions though, it is necessary to confirm this finding in other cohorts. Nevertheless, the oxygen-based endotypes still trended as expected in most cases, providing reassurance that they are capturing the physiological processes that they are intended to measure.

The graphical endotype model was verified by comparing the gap—defined as the difference between the SpO2 arousal threshold and the steady-state SpO2—with various indicators of OSA severity. This comparison revealed a robust correlation between the gap and OSA severity, irrespective of the severity metric employed. This association provides evidence that the model agrees with physiological expectations.

While we would like to compare the oxygen-based endotypes with the PSG-derived endotypes (5–7) in this cohort, the latter require nasal pressure measurements, which were not performed in the SHHS. Moreover, the endotypes represent slightly different physiological processes in the two methods. In the PSG method, there is a separate determination of loop gain, whereas in the oxygen-based method the loop gain components—controller gain and plant gain—are wrapped into the SpO2 arousal threshold and steady-state SpO2 endotypes, respectively. Furthermore, the oxygen-based method culminates in a model that demonstrates the interplay of the endotypes in producing OSA, a feature not inherently provided by the PSG method. These differences complicate direct comparisons between the two approaches.

Overall, we believe the findings from the current study represent an important step forward in the endotyping of patients with OSA, offering a novel approach that overcomes some of the hurdles encountered in previous endotyping methods.

### Plant gain for SaO2

As mentioned above, the steady-state SpO2 is influenced by plant gain, defined here as the change in SaO2 for a given change in ventilation. Figure 6 illustrates the plant gain, with the normal condition represented by the solid blue line. Plant gain is nonlinear: steep during hypoventilation and flat during hyperventilation. As a result, hypoventilation raises plant gain, whereas hyperventilation lowers it. Low barometric pressure and V/Q mismatch also increase plant gain. V/Q mismatch (dot-dash line in Figure 6) elevates plant gain because low V/Q units operate on the steep part of the oxyhemoglobin curve. Interestingly, shunt (dotted line) does not affect plant gain because, even though shunted units are on the steep portion of the oxyhemoglobin curve, there is no ventilation to remove from them.

**Figure 6.**
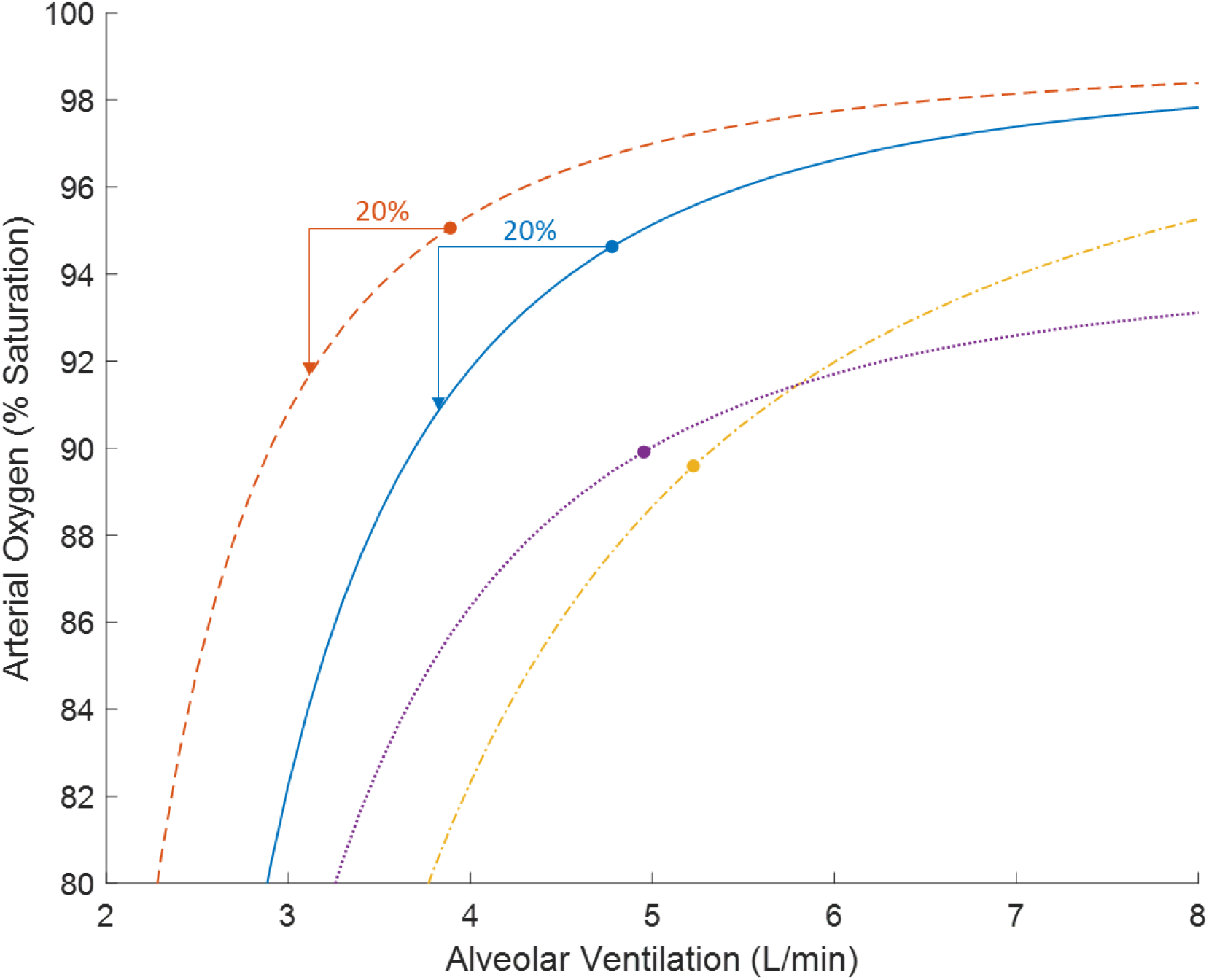
Steady-state plant gain for SaO2 under normal conditions (solid blue line), reduced metabolic rate (dashed orange line), ventilation-perfusion mismatch (dot-dash yellow line), and shunt (dotted purple line). The circles represent baseline SaO2 and ventilation.

Metabolic rate also influences plant gain. A reduced metabolic rate shifts the plant gain line left, lowering ventilation but increasing baseline SaO2 due to reduced oxygen consumption. However, this effect disappears when the ventilatory reduction is expressed as a percentage decrease rather than an absolute reduction (shown by the arrows in Figure 6).

Lung volume itself does not affect steady-state plant gain but influences how quickly SaO2 reaches a steady state. Obesity impacts lung volume (25–29) but does not cause ventilation heterogeneities until severe enough to nearly eliminate expiratory reserve volume (30–33). These individuals would likely have a low awake SpO2, making them poor candidates for the oxygen-based endotyping method.

Low cardiac output can indirectly increase plant gain through pulmonary congestion and V/Q mismatch. However, it does not affect steady-state plant gain if enough time has passed for blood recirculation to stabilize. More importantly, reduced cardiac output is often associated with central apneas, which can confound the interpretation of the steady-state SpO2 endotype. Therefore, the oxygen-based endotyping method is likely not valid in patients with congestive heart failure.

In summary, the factors affecting steady-state plant gain for SaO2 are those that produce hypoxemia, namely hypoventilation, V/Q mismatch, and low barometric pressure. In patients with these conditions, steady-state SpO2 should be interpreted cautiously because it reflects not only the reduction in ventilation but also the elevated plant gain.

### SpO2 as an arousal threshold metric

Research indicates that hypoxia is a weak arousal stimulus when PCO2 levels are normal or low (34–36). In hyperoxic conditions, PCO2 does not induce arousal until it rises about 15 mmHg above baseline (18, 35, 37, 38). At a controller gain of 1.5 L/min/mmHg, this corresponds to a ventilatory drive increase of 23 L/min, suggesting that hypercapnia alone is also a weak arousal stimulus. However, during upper airway obstruction, hypoxia and hypercapnia occur together and have a synergistic effect on ventilatory drive (39). Therefore, it is important to consider both gases when assessing how chemical stimuli produce arousal.

More critically for our purposes, the peripheral chemoreceptors are sensitive to hypoxia (40, 41) and play an important role in arousals (42, 43). Moreover, due to their rapid response time (44, 45), peripheral chemoreceptors are the predominant regulators of ventilation during transient apneas (17). While the central chemoreceptors are twice as strong as their peripheral counterparts (46), their response time of 2 to 3 minutes (44, 45) renders them less effective in responding to gas tensions that cycle every 30 seconds. Since the peripheral chemoreceptors are sensitive to hypoxia and are the primary controllers of ventilation during transient apneas, we believe SpO2 is a reasonable arousal threshold metric.

It is important to note that, due to the circulatory delay, the oxygen level sensed by the chemoreceptors at the time of arousal is higher than the nadir SpO2. While this could be corrected by assuming a 6-second delay (47), we ultimately chose not to do this because the nadir is easier to identify in an all-night tracing, thereby allowing statistical estimates to be visually confirmed. Nevertheless, it is worth exploring whether to make such a correction in future iterations of this method.

## Conclusions

In conclusion, this study demonstrates that the proposed oxygen-based method for endotyping OSA is feasible and aligns with physiological expectations in most cases, as indicated by the anticipated changes in endotypes with body position and sleep stage. While these results are encouraging, further validation is needed to understand how these endotypes change with treatment and to assess the model’s ability to predict treatment outcomes. This additional research will be crucial in determining the clinical utility of this method for guiding personalized therapeutic strategies for OSA patients.

## Data Availability

All data produced in the present study are available upon reasonable request to the authors

## Acknowledgements

none.

## Impact

Oxygen-based endotypes of Obstructive Sleep Apnea (OSA) could impact both clinical medicine and basic science. Clinically, the ability to measure endotypes using pulse oximetry makes it more accessible and less invasive, potentially improving diagnostic accuracy and personalized treatment plans. In basic science, this method provides a deeper understanding of the physiological mechanisms underlying OSA, adding to knowledge of the disease process and aiding the development of targeted therapies.

## Author’s contributions

Conception: AW. Study design: AW. Data analysis: AW, LM, Ali Azarbarzin, NE, SS. Statistical analysis: AW. Manuscript Draft: AW. All authors interpreted data, edited the manuscript for important intellectual content, and approved the final draft.

